# Racial and Socioeconomic Disparities in ICU Admissions Among Obstetric Patients at a Tertiary Urban Center

**DOI:** 10.64898/2026.04.04.25343104

**Authors:** Valerie Martin

## Abstract

We aimed to evaluate disparities in perinatal ICU admissions at an urban medical center and to contextualize these findings relative to national U.S. data provided by the Centers for Disease Control and Prevention (CDC). To do so, we performed a retrospective review of all pregnant and < 6-week postpartum patients admitted to the ICU between October 2023 and June 2025. The cohort included 58 patients: 81% were non-Hispanic Black, and 91% were publicly insured. These local data can be compared to national data, which demonstrate higher rates of severe maternal morbidity (SMM) and ICU admission among Black patients and those insured by Medicaid. In 2023, the U.S. maternal mortality rate was 18.6 per 100,000 live births, down from 22.3 in 2022. However, significant disparities persist, with mortality rates of 50.3 per 100,000 among Black women compared with 14.5 per 100,000 among White women[1]. The most frequently reported indications for obstetric ICU admission include hypertensive disorders of pregnancy, obstetric hemorrhage, and severe underlying medical comorbidities[9,10].

## Introduction

Severe maternal morbidity (SMM) and mortality remain inappropriately high in the United States and are characterized by persistent racial and socioeconomic inequities. Building on national data, prior literature suggests that observed disparities are driven by structural and systemic factors, including limited access to prenatal care, cumulative physiologic effects of chronic stress associated with racism, and inequities in how care is delivered[4,6]. According to the American College of Obstetricians and Gynecologists, strategies to address these inequities include implementation of standardized obstetric early warning protocols, routine multidisciplinary review of severe maternal morbidity and ICU admissions, and enhanced integration with community-based resources[1].

## Methods

We conducted a retrospective review of pregnant or ≤6-week postpartum patients admitted to the ICU at a tertiary urban hospital from October 2023 to June 2025 (N=58). We collected data on age, registered race/ethnicity, insurance status, medical comorbidities, and critical interventions such as cesarean delivery, hysterectomy, or embolization. To further analyze disparities, patients were categorized by race/ethnicity (Black/African American, Hispanic/Latina, Caucasian) and payer type (Medicaid/public, private commercial, uninsured/charity). Descriptive statistics summarized demographics and indications. To contextualize findings, we compared results with published U.S. reports and national maternal outcome data taken from the CDC vital statistics website[1,6]. Census data from the American Community Survey(ACS) for the hospital’s zip code were used as control benchmarks for the cohort.

## Results

Among 58 obstetric ICU admissions, 47 (81%) were Black/African American and 7 (12%) were Hispanic/Latina. The remaining 7% were Caucasian. In terms of payer status, 53 (91%) had Medicaid or charity care, and 4 (7%) had commercial insurance. The median maternal age was 30 years. These demographic findings set the stage for a closer look at the reasons for ICU admission and how they align with national patterns.

The most common primary ICU indications reflected national trends. Specifically, hypertensive disorders of pregnancy, including severe preeclampsia, eclampsia, and HELLP syndrome, accounted for about 38% of cases. Obstetric hemorrhage, including placental abruption and placenta accreta spectrum, accounted for 17%, and major medical comorbidities such as cardiac, respiratory, or neurologic events accounted for 24%. Psychiatric or substance-related admissions comprised of 9%. Many patients had multiple diagnoses, but hypertension and bleeding were most prevalent. Nearly 60% of the cohort were delivered during their ICU hospitalization. Cesarean delivery occurred in 48%, operative vaginal delivery in 7%, and hysterectomy or uterine artery embolization in 10%. Five patients (9%) experienced ICU cardiac arrest or cardiopulmonary collapse. Together, these characteristics illustrate the overlap between patient complexity and severe outcomes.

Black and publicly insured patients were overrepresented in the ICU cohort compared to the base population. For reference, Census/ACS data for the hospital’s zip code showed that 84.5% of residents were Black. More notably, only 51.5% of the community has private insurance, yet 91% of ICU patients had Medicaid or charity care. Only 7% had commercial insurance, compared to 51.5% in the community, indicating a significant underrepresentation. The community uninsured rate is 14.2%, but many uninsured women likely enter pregnancy without coverage and rely on charity care, contributing to the high proportion of publicly insured ICU patients. These contrasts highlight that ICU admissions at this center disproportionately involved women from the most socioeconomically vulnerable and minority groups, consistent with previous reports[3,4].

Nationally, Black women face a significantly higher burden of severe maternal complications. As highlighted, CDC data show that in 2023, the U.S. maternal mortality rate decreased to 18.6 per 100,000 live births (from 22.3 in 2022), but the racial gap persists: 50.3 per 100,000 for Black women compared to 14.5 for White women. □ Previous literature indicates that Black patients are two to three times more likely than White patients to experience life-threatening obstetric events requiring ICU care, even after adjusting for age and comorbidities[3,4]. Medicaid insurance is also an independent risk factor, with maternal ICU admission rates in Medicaid populations often more than double those in privately insured groups.^1^□ The local findings (81% Black, 91% Medicaid ICU patients) align with these national SMM trends, highlighting the close association between critical maternal illness, race, and insurance status. This connection underscores the broader impact of these disparities at both the national and local levels.

## Discussion

The cohort’s demographic profile reflects institutional disparities. Importantly, race and insurance serve as social constructs that correlate with underlying determinants. Black women in the United States experience higher rates of chronic stress, hypertension, and structural disadvantage, predisposing them to severe preeclampsia and cardiomyopathy. Furthermore, public insurance is often associated with fragmented prenatal care, transportation barriers, and reduced access to high-risk perinatal specialists. These factors likely contributed to the critical conditions observed in the cohort. More than half of the patients had identifiable high-risk features, such as multiple prior Cesareans with morbidly adherent placenta, sickle cell disease, heart disease, or severe substance use, which are known to occur at higher prevalence in at-risk communities. The convergence of these factors illustrates the systemic contributors to severe maternal outcomes.

Race is not a biological cause of severe illness, but a proxy for differential exposure to risks such as racism, poverty, and environmental stress. Both SMFM and ACOG accentuate that disparities result from structural racism and social determinants, including housing, education, and occupational exposures, rather than innate racial differences[1,8]. These principles are reflected in our findings: Black patients disproportionately require ICU care due to unequal access and cumulative burdens, not immutable traits. For example, patients from low-income neighborhoods may delay prenatal visits until hypertensive crises become severe. Additionally, implicit bias may also delay escalation of care, as studies suggest minority women often receive ICU transfers at higher acuity thresholds[4]. Understanding these contributing factors is essential for designing effective interventions.

These disparities necessitate an equity-centered quality improvement approach. As a first step, three key strategies are recommended: a multidisciplinary review of each obstetric ICU admission (and any SMM event) should be implemented, explicitly addressing race and insurance factors. For example, the assessment should include whether prenatal care was initiated early and whether social needs were screened. SMFM recommends including equity metrics in SMM reviews, reinforcing the importance of these considerations in practice[7].

- Standardized Early Warning and Escalation: Next, develop and implement uniform obstetric early warning systems for hypertension and hemorrhage. Use standardized blood pressure protocols (such as ACOG’s BP management guidelines) and hemorrhage checklists with clear ICU triggers to reduce subjective variation. This consistent approach can mitigate variations in care escalation[2].
- Community and Social Supports: Finally, strengthen connections to community resources, including social work, transportation programs, and chronic disease management, for high-risk patients. Given the link between public insurance and ICU risk, hospitals should collaborate with Medicaid programs to identify and enroll at-risk pregnant women early. Together, these steps form a coordinated response to address disparities beyond hospital walls.

The proposed process begins with prenatal risk screening (such as a checklist), leading to intensified monitoring or antepartum admission for hypertensive or hemorrhagic risk. If deterioration occurs, a protocol-driven escalation, such as activating an obstetric rapid response team or ICU consult, might be triggered. Each ICU admission is then reviewed by a root-cause team that includes an “equity champion” to address community factors, ensuring that every step is linked to an equity-oriented review.

This retrospective single-center study has limited generalizability and does not include a denominator of all deliveries for rate calculations. The small sample size prevented complex data analysis. Race and insurance data were obtained from EMR registration, which may contain inaccuracies. However, the results closely mirror known disparities. To improve validity, future research should link ICU cases to the full institutional delivery cohort to calculate ICU admission rates by race and insurance, and could use more complex data evaluation models to identify meaningful interpretations. Interviews with patients and providers may clarify case-specific barriers, such as delays in seeking care, and help optimize resources for patients.

In summary, these findings emphasize that obstetric ICU admissions are not randomly distributed, but instead disproportionately affect patients experiencing structural and socioeconomic disadvantages. The overrepresentation of non-Hispanic Black and publicly insured patients in ICU admissions at this institution mirrors national patterns and highlights persistent gaps in access, care delivery, and health system responsiveness. These disparities impact maternal morbidity and mortality and represent a critical opportunity for intervention. Implementing standardized clinical protocols, equity-focused review processes, and strengthening community partnerships are recommended to improve outcomes. Addressing these inequities is fundamental not only to advancing maternal health but to achieving a more equitable healthcare system.

**Table 1.**
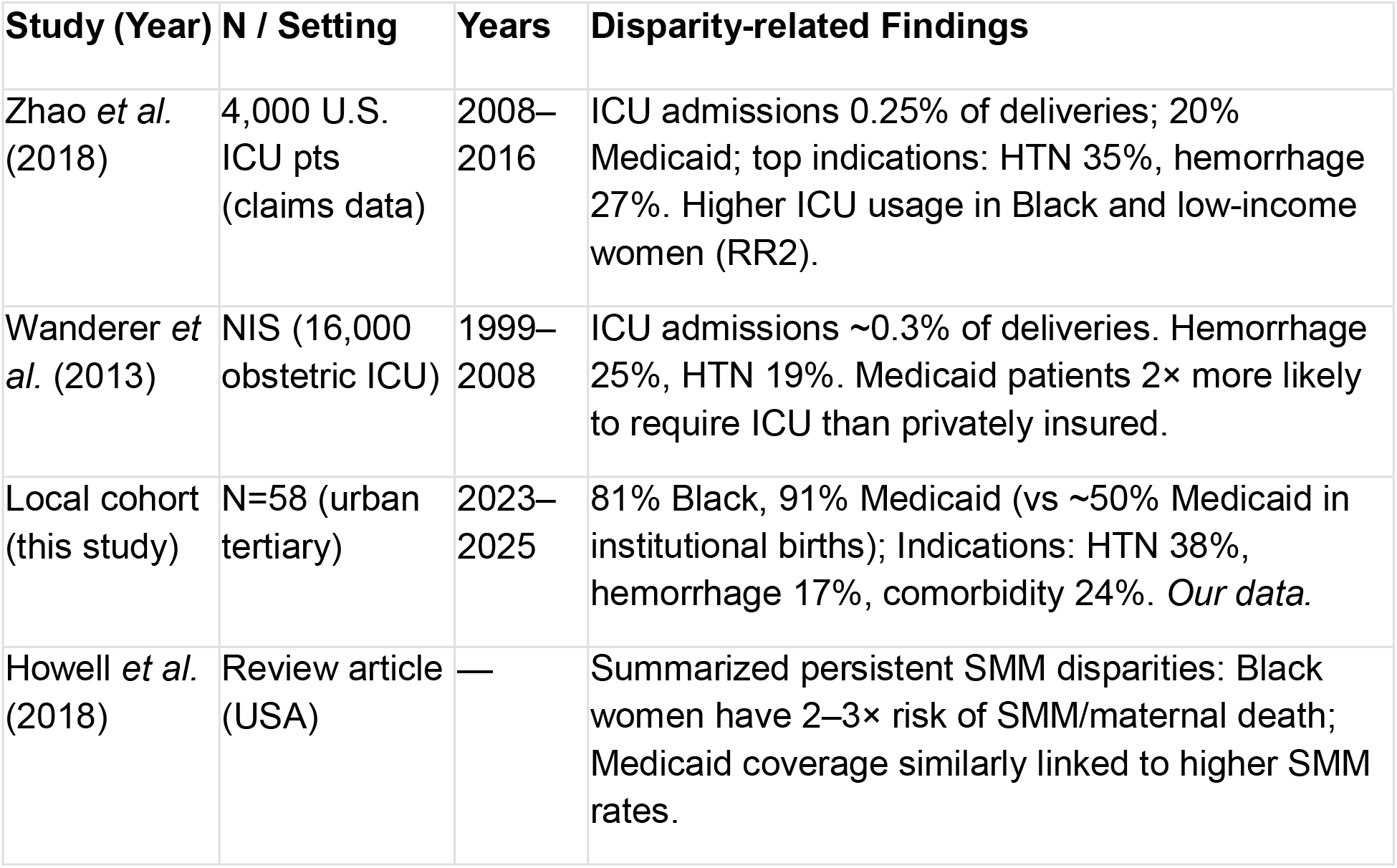
Key recent studies of obstetric ICU admissions and disparities.

| Study (Year) | N / Setting | Years | Disparity-related Findings |
| --- | --- | --- | --- |
| Zhao *et al.* (2018) | 4,000 U.S. ICU pts (claims data) | 2008–2016 | ICU admissions 0.25% of deliveries; 20% Medicaid; top indications: HTN 35%, hemorrhage 27%. Higher ICU usage in Black and low-income women (RR2). |
| Wanderer *et al.* (2013) | NIS (16,000 obstetric ICU) | 1999–2008 | ICU admissions ~0.3% of deliveries. Hemorrhage 25%, HTN 19%. Medicaid patients 2x more likely to require ICU than privately insured. |
| Local cohort (this study) | N=58 (urban tertiary) | 2023–2025 | 81% Black, 91% Medicaid (vs ~50% Medicaid in institutional births); Indications: HTN 38%, hemorrhage 17%, comorbidity 24%. *Our data.* |
| Howell *et al.* (2018) | Review article (USA) | — | Summarized persistent SMM disparities: Black women have 2–3x risk of SMM/maternal death; Medicaid coverage similarly linked to higher SMM rates. |

## Data Availability

All data produced in the present study are available upon reasonable request to the authors

## Notes

### Competing Interest Statement

The authors have declared no competing interest.

### Funding Statement

This study did not receive any funding.

### Author Declarations

Institutional Review Board at Newark Beth Israel waived ethical approval for this work.

## References

1. American College of Obstetricians and Gynecologists; Society for Maternal-Fetal Medicine. Obstetric care consensus No. 5: severe maternal morbidity: screening and review. Obstet Gynecol. 2016;128(3):e54–e60. doi:10.1097/AOG.0000000000001642

2. American College of Obstetricians and Gynecologists. ACOG practice bulletin No. 211: critical care in pregnancy. Obstet Gynecol. 2019;133(5):e303–e319.

3. Clark SL, et al. Severe maternal morbidity and mortality: disparities in outcomes. Obstet Gynecol. 2017;129(5):781–789.

4. Howell EA. Reducing disparities in severe maternal morbidity and mortality. Clin Obstet Gynecol. 2018;61(2):387–399.

5. Hoyert DL. Maternal mortality rates in the United States, 2023. National Center for Health Statistics; Centers for Disease Control and Prevention. 2024.

6. Petersen EE, Davis NL, Goodman D, et al. Racial/ethnic disparities in pregnancy-related deaths—United States, 2007–2016. MMWR Morb Mortal Wkly Rep. 2019;68(35):762–765.

7. Society for Maternal-Fetal Medicine. SMFM clinical consult series: addressing systemic racism in obstetrics. Published 2020.

8. Society for Maternal-Fetal Medicine. SMFM special statement: the use of race in maternal-fetal medicine research. Published 2021.

9. Wanderer JP, Leffert LR, Mhyre JM, Kuklina EV, Callaghan WM, Bateman BT. Epidemiology of obstetric-related ICU admissions in the United States. Anesthesiology. 2013;118(5):1074–1083.

10. Zhao Y, et al. Trends and characteristics of obstetric ICU admissions in the United States. Crit Care Med. 2018;46(9):1466–1472.

